# SROTAS IQ: An AI-Based Clinical Trial Matching Platform: A Validation Study in Breast Cancer

**DOI:** 10.1101/2025.08.13.25333342

**Authors:** Samuel McInerney, Heeba Gurku, Ramji Balasubramanian, Vikram Parimi, Suman Bhaskaran, Karthik Sekaran

## Abstract

**Objectives:** To evaluate the performance of SROTAS IQ, a custom fine-tuned large language model (LLM), in automating clinical trial eligibility screening for breast cancer patients using synthetic data.

**Methods:** Ten breast cancer trials were selected across diverse treatment settings and molecular subtypes. Fifteen synthetic patient summaries per trial were generated, including realistic and enriched eligibility scenarios. Two independent oncologists assessed trial eligibility for each patient, establishing ground truth. SROTAS IQ LLM was evaluated against expert consensus using standard classification metrics. Time-to-verdict was measured to compare clinician effort with automated assessment.

**Results:** SROTAS IQ demonstrated strong concordance with expert assessments, achieving 90% or greater accuracy in 5 of 10 trials. Across 150 patient-trial evaluations, the model correctly classified 88% of overall eligibility decisions. Performance was highest in trials with moderate complexity and fewer nested criteria, while more intricate protocols showed reduced accuracy. The LLM consistently delivered rapid assessments (<0.5 minutes per patient), with explainable outputs that aligned with clinical reasoning. These findings underscore the model’s potential to support high-fidelity, scalable trial matching in oncology.

**Conclusion:** SROTAS IQ offers a promising approach to automating clinical trial matching in oncology. Further real-world validation is needed to confirm generalisability and integration into clinical practice.

## 1. Introduction

The landscape of cancer clinical trials has evolved dramatically over the past decade, with increasing emphasis on precision medicine approaches that require sophisticated biomarker-driven patient selection [1]. While this evolution has led to more targeted and potentially effective therapies, it created significant challenges in identifying appropriate trial opportunities for individual patients [2]. Modern Phase III oncology trials often include 30 to 40 distinct inclusion and exclusion criteria, and some biomarker-driven protocols exceed 50 requirements spanning laboratory values, imaging findings, treatment history, performance status, and molecular characteristics [3]. This complexity, combined with practical constraints in clinical workflows, creates substantial barriers to optimal trial recruitment.

Disparities in trial access persist, particularly for patients in community settings or with complex comorbidities. Manual matching processes may inadvertently exclude eligible individuals, underscoring the need for scalable, equitable solutions. [4][5].

Large language models (LLMs) offer a promising alternative, capable of interpreting nuanced clinical documentation and synthesizing information across multiple data sources. [6]. Unlike rule-based systems, LLMs can understand temporal relationships and contextual dependencies, enabling more accurate eligibility assessments.

Recent models such as PRISM, OncoLLM [8], and TrialGPT [6] have demonstrated potential in automating trial matching. However, challenges remain in handling complex trial protocols, diverse disease subtypes, and edge cases. Performance often declines with increasing trial complexity, and infrastructure demands can be substantial.

This study evaluates the clinical accuracy of SROTAS IQ, a custom fine-tuned LLM designed for breast cancer trial matching. Using synthetic patient data and expert oncologist validation, we assess the model’s ability to identify eligible candidates across diverse trial scenarios. Key metrics include matching accuracy, explainability, and time-to-verdict.

We assessed the effectiveness and performance of the custom domain-specific LLM (SROTAS IQ LLM) against expert oncologist evaluation, aiming to harness the potential of generative AI in simplifying the complex patient–clinical trial matching process. Our primary focus was on eligibility classification accuracy across diverse synthetic patient scenarios and trial types. In addition, time-to-verdict was measured to illustrate potential efficiency gains, comparing the time taken by clinicians and the LLM to assess trial eligibility. This comparison offers insight into the effort required for manual versus automated review, particularly as the complexity of inclusion/exclusion criteria and patient data increases.

We present a comprehensive methodology encompassing synthetic patient data generation, expert validation, and model evaluation. The results demonstrate promising performance and clinical coherence, with implications for future deployment, limitations, and areas for further research.

## 2. Methods

We consider the task of automated patient-to-trial matching for clinical trial eligibility assessment. *P* = {*p*_*1*_, *p*_*2*_, …*p*_*n*_}, denotes a set of synthetic patient information and *T* = {*t*_*1*_, *t*_*2*_, …*t*_*m*_} as a set of clinical trials.

Each trial t_i_ □ *T* is defined by a set of eligibility criteria from the trial information, 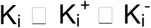 where, 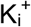 and K_i_ represent inclusion and exclusion criteria, respectively. For each patient p_i_ □ P, trial t_i_ □ *T*, the eligibility of the patient p_i_ is assessed based on each criterion k □ Ki. The outcome for each criterion is denoted as a categorical label l □ □, where □ = {*Met, Not Met, Unknown*}. The expert annotated ground truth is labelled as ε: *P * T * K −>* □, where ε(p_i_,t_i_, k) = l. The ground truth serves as a reference standard to perform model evaluation.

We constructed a synthetic training dataset *D* containing samples consisting of trial and labelled patient information. The large language model *M* is trained over a dataset of size 2707 to predict the eligibility. During inferencing, for each trial, t_i_ □ *T*, with a set of criteria K_i_ and 15 synthetic patients are evaluated with *M*, where y’ = *M*(prompt (p_i_, K_i_).

### 2.1. Dataset Characteristics

A team of oncology specialists generated 2707 synthetic breast cancer patient summaries representing diverse clinical scenarios, including early-stage, locally advanced, and metastatic disease. Molecular subtypes included hormone receptor-positive, HER2-positive, and triple-negative cases. Summaries were derived from open-access case reports and validated by oncologists for clinical coherence.

Additionally, summaries were extracted from breast cancer case reports in PubMed Central (PMC), an open-access repository. These cases enhanced clinical diversity and introduced natural linguistic variation into the dataset [9].

Patient demographics reflected real-world breast cancer populations with a mean age of 56.5 years (range: 34-79 years) and an appropriate distribution of performance status, comorbidity patterns, and treatment history complexity. Clinical coherence validation by expert oncologists confirmed that synthetic patient scenarios accurately reflected real-world clinical complexity while maintaining controlled characteristics necessary for systematic evaluation. The dataset successfully included challenging edge cases representing a significant number of the total cohort, including patients with borderline performance status, complex treatment histories, and significant comorbidities that impact trial eligibility.

### 2.2 Validation Dataset Generation

For the validation study, we generated 15 synthetic breast cancer patient cases based on realistic clinical scenarios to assess clinical trial matching performance. These validation patients were created independently from the training dataset to ensure unbiased evaluation of system performance. The validation dataset consisted of 10 patients designed to include realistic eligibility distributions rather than perfect matches, and the 5 enriched patients were designed to meet more eligibility criteria, ensuring an adequate sample of both positive and negative matches for model testing.

Both sets incorporated clinical complexity and documentation variability typical of real-world oncology practice. The proprietary AI system Claude Sonnet 4 was used to generate these summaries with the prompts provided in **Supplementary File 1**

Each synthetic patient summary underwent rigorous clinical coherence validation through expert review by medical oncologists who assessed the internal consistency of clinical scenarios, appropriateness of treatment sequences, realistic biomarker and staging combinations, and overall clinical plausibility. This validation process ensured that synthetic cases accurately reflected real-world clinical complexity while maintaining the controlled characteristics necessary for systematic evaluation.

### 2.3 Clinical Trial Selection

We selected breast cancer trials from ClinicalTrials.gov (**https://clinicaltrials.gov/**) using systematic search criteria designed to represent major clinical contexts. We extracted trials across three key dimensions: 1. treatment setting (neo-adjuvant, adjuvant, metastatic) 2. molecular subtypes (hormone receptor positive/negative, HER2 positive/negative) and 3. Disease stages (Early, locally advanced, metastatic). Trials were focused on systemic anti-cancer interventions rather than radiotherapy or surgical interventions.

From these comprehensive search results, we selected a focused set of 10 trials that provided representative coverage across treatment paradigms while maintaining practical feasibility for expert validation. Trial complexity varied across the dataset, with some studies containing 10-15 eligibility criteria and a few scenarios with complex biomarker-driven or combination trials containing 20-30 or more criteria. The systematic extraction and selection approach enabled comprehensive coverage of contemporary breast cancer trial landscape while ensuring a manageable scope for rigorous expert review. Each trial’s eligibility criteria were extracted and categorised to enable analysis of system performance across different trial types and complexity levels.

### 2.4 Clinician Validation

The system was evaluated by two oncology specialists with substantial experience in breast cancer treatment and clinical trial management. Each expert independently reviewed all synthetic patient cases and provided trial-matching recommendations without knowledge of the LLM system outputs or the other reviewer’s assessments, ensuring unbiased ground truth establishment. The two-expert approach was selected to balance practical feasibility with robust validation, providing sufficient clinical expertise while remaining manageable for comprehensive case review.

Each oncologist evaluated 150 patient-trial combinations (10 trials × 15 patients per trial) using a ternary assessment system: Met, Not Met, or Unknown for each eligibility criterion, followed by an overall trial eligibility decision. For cases with disagreement between the two oncologists, structured discussion was conducted to reach consensus or flag cases as inherently ambiguous scenarios highlighting areas of legitimate clinical uncertainty.

Inter-rater reliability was assessed using Cohen’s kappa. For cases with disagreement, oncologists discussed and reached consensus decisions, which were then used as ground truth to evaluate SROTAS IQ LLM accuracy.

### 2.5. SrotasIQ LLM Performance Evaluation

SROTAS IQ LLM performance was evaluated using standard classification metrics comparing system outputs to expert consensus decisions. For each patient-trial combination, individual eligibility criteria were assessed using a ternary classification system (Met/Not Met/Unknown), followed by an overall trial eligibility determination.

Performance Metrics: Primary metrics included sensitivity (true positive rate), specificity (true negative rate), positive predictive value (PPV), negative predictive value (NPV), precision, recall, and F1 score. These were calculated for each patient-trial combination based on individual criteria assessments, with overall accuracy determined by the proportion of correctly assessed criteria out of total criteria per trial.

Handling of “Unknown” Responses: Special consideration was given to LLM “Unknown” assessments. The system tended to mark criteria as “Unknown” when information was not explicitly stated, whereas clinicians typically inferred “Not Met” for unstated criteria. “Unknown” responses were evaluated for clinical appropriateness, considered correct when information was genuinely absent but incorrect when sufficient information existed for reasonable clinical inference.

In the case of an unknown LLM response when there was a different answer provided by the ground truth, the clinical reasoning provided by the SROTAS IQ LLM was reviewed alongside the synthetic patient history. A decision was then made by the oncologist as to whether this was a reasonable inference.

Statistical analysis included calculation of inter-rater reliability using Cohen’s kappa, confidence interval estimation for primary performance metrics, stratified analysis examining performance across patient subgroups (realistic vs enriched cases) and trial complexity categories, and error pattern analysis to identify systematic biases and guide system improvements.

To assess the comparative effort required for clinical trial eligibility screening, we measured the time taken by expert oncologists versus the SROTAS IQ LLM across all patient-trial evaluations. Oncologist screening times were recorded manually, while LLM assessments were computed automatically.

In real-world settings, eligibility screening often involves navigating multiple systems to retrieve lab results, imaging reports, and treatment histories. In contrast, our synthetic patient summaries were concise and self-contained, reducing the cognitive and logistical burden on clinicians. As a result, the average time per patient for oncologist evaluation was lower than typical clinical scenarios.

We observed a learning curve among oncologists: initial evaluations took 15–25 minutes per patient, decreasing to 5–10 minutes as reviewers became familiar with the trial protocols and summary format. This adaptation reflects the efficiency gains possible with structured, disease-specific summaries—in this case, tailored to breast cancer.

Importantly, the SROTAS IQ LLM completed each assessment in under 0.5 minutes, demonstrating substantial time savings. While this comparison is constrained by the synthetic nature of the data and controlled presentation format, it highlights the potential for AI-assisted screening to reduce clinician workload and accelerate trial recruitment workflows.

### 2.6. SROTAS IQ *LLM*

SROTAS IQ LLM is a custom fine-tuned model, trained on a wide category of synthetic breast cancer patient data. It effectively interprets and decodes complex eligibility criteria that often contain nested conditions, time-dependent requirements, and context-dependent exclusions. The LLM generates matching recommendations and delivers relevant patient information suitable for specific trials. It is accompanied by detailed explanations, provides transparent reasoning for inclusion and exclusion decisions based on the evaluation of the criteria. This explanatory component was specifically designed to support clinicians during clinical decision-making, enabling physicians to quickly understand, interpret, and validate the system’s reasoning and recommendations. We used Mistral 7B (v0.3) large language model, to build the SROTAS IQ LLM. It’s adopted due to its commendable performance over the benchmark state-of-the-art models. SROTAS IQ LLM was constructed by fine-tuning Mistral-7B with Low-Rank Adaptation (LoRA) [10]. We ensured the model parameters are designed to efficiently capture the domain-specific understanding from the training process. The LoRA parameters are dropout (0.01) and rank (64). The model fine-tuning was carried out on Azure ML through a managed GPU cluster with an NVIDIA A100.

## 3. Results

The clinical trial patient eligibility assessment evaluation between the clinician and SROTAS IQ LLM is performed to determine the LLM’s ability. The results are compared and validated by the two independent clinicians. The clinician’s evaluation of each trial criterion against the patient summary is captured individually. The mismatch between the clinician’s opinion is addressed by introducing a consensus.

Each oncologist independently evaluated 150 patient-trial combinations, with subsequent consensus discussion for discordant cases. Inter-rater reliability between the two oncologists was good (Cohen’s κ = 0.742, 95% CI: 0.48-0.89). With 84.7% overall agreement across all 150 patient-trial combinations before consensus discussion. Time-to-verdict is calculated to compare the actual time difference between human evaluation and SROTAS IQ LLM.

Furthermore, this ground-truth is compared to the responses generated by the SROTAS IQ LLM for each trial criterion. This experiment is conducted across all 10 clinical trials, each involving 15 individual patients. The study’s results highlight that the custom model performs at a level comparable to human experts in clinical efficacy. The scores for each trial group are displayed as a radar chart, shown in Figure 2. It visualizes the model’s performance across various metrics for each patient. The values are obtained by averaging the scores of all patients within each trial, with each axis representing a different metric. The consistency and stability are indicated by the uniformity in shape and size across the patients.

**Figure 1.**
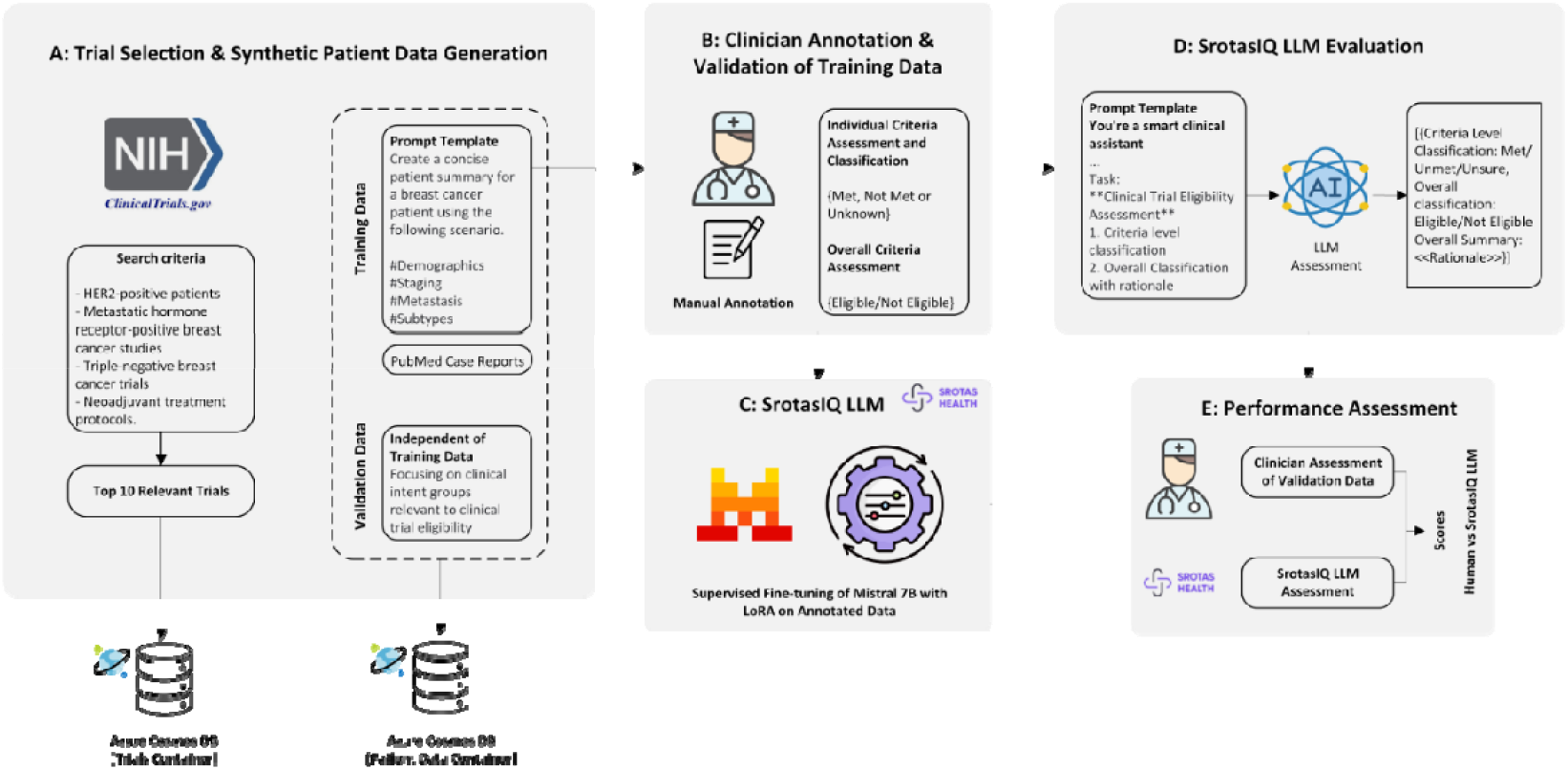
Workflow of the overall proposed system. Each process is categorised into modules from A to E. Module A describes the trial selection & synthetic patient data generation performed by a team of oncologists. The clinical trial information, as well as the training and validation datasets are stored in Azure Cosmos DB. In Module B, the clinical annotation and validation of the training data are carried out by the oncologists. Module C involves the preparation of the SROTAS IQ LLM, custom trained on the synthetic training dataset on Mistral 7B (v0.3) model using LoRA. Module 4 discusses the evaluation of SROTAS IQ LLM on the validation dataset. In Module 5, the performance of the SROTAS IQ LLM against the oncologists’ annotated results is compared and assessed through various metrics.

**Figure 2.**
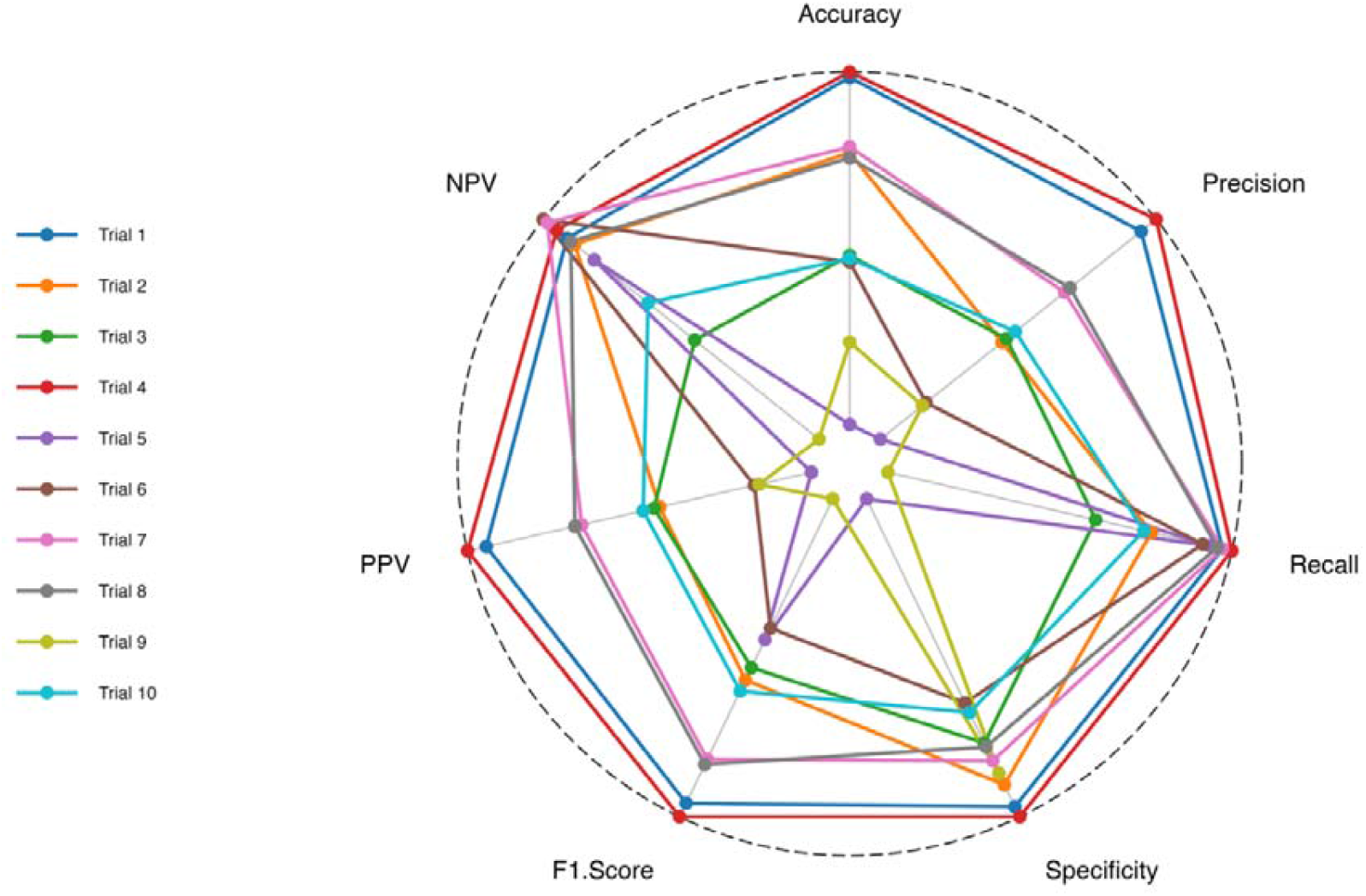
Radar chart representing the performance of SrotasIQ LLM across the trials 1 to 10 evaluated under different metrics. The data used to plot the chart is normalized with min-max technique to increase the visibility and showcase the difference in performance among the trials.

The min-max normalisation technique was applied to the actual data, which improved the visibility and differentiation of the performance among trials against various evaluation metrics. The actual and the normalised scores of the trials with metrics are provided in **Supplementary File 2**. From the radar chart, the LLM performance of Trials 1, 2, 4, 7 and 8 is the most accurate, whereas Trials 3, 5, 6, 9 and 10 had a poor classification of the criteria. Apart from Trial 5, all the remaining were contained more than 23 criteria, with an average of 26.6 in the poor classification group. Conversely, only an average of 20.8 criteria in the other category, with 27 criteria in Trial 1 as the highest. This observation is important to understand the model’s behaviour with varying criteria size in each trial. Moreover, this difference in the results is a good indicator that the model is not overfit to the patient data with the trial criteria. Despite the normalised representation of the radar chart, the actual results across the trials are tabulated in Table 1.

**Table 1:** Average performance of SrotasIQ LLM across 10 trials evaluated with various metrics.

| Trial | Criteria | Accuracy | Precision | Recall | Specificity | F1-Score | PPV | NPV |
| --- | --- | --- | --- | --- | --- | --- | --- | --- |
| Trial 1 | 27 | 97.79 | 95.83 | 97.35 | 96.65 | 96.37 | 95.83 | 97.77 |
| Trial 2 | 20 | 92.96 | 79.47 | 88.11 | 94.13 | 81.21 | 79.47 | 97.32 |
| Trial 3 | 24 | 86.38 | 79.99 | 80.85 | 89.32 | 79.69 | 79.99 | 90.23 |
| Trial 4 | 20 | 98.14 | 97.60 | 98.56 | 97.82 | 98.04 | 97.60 | 98.38 |
| Trial 5 | 15 | 75.55 | 65.15 | 95.66 | 61.17 | 76.23 | 65.15 | 96.16 |
| Trial 6 | 28 | 85.95 | 70.53 | 94.77 | 84.70 | 74.82 | 70.53 | 99.17 |
| Trial 7 | 18 | 93.33 | 86.83 | 97.33 | 91.36 | 90.97 | 86.83 | 98.93 |
| Trial 8 | 19 | 92.63 | 87.47 | 96.66 | 89.76 | 91.60 | 87.47 | 97.54 |
| Trial 9 | 38 | 80.83 | 70.10 | 53.83 | 92.81 | 58.91 | 70.10 | 82.89 |
| Trial 10 | 28 | 86.19 | 81.03 | 87.12 | 85.80 | 82.56 | 81.03 | 92.98 |

The heatmap in Figure 3 depicts the overall trial eligibility classification predicted by the LLM and its evaluation against the human expert. The eligibility could either be “Eligible” or “Not Eligible”. The colour gradients represent the match/noMatch, where green indicates the correct match between human expert evaluation and LLM prediction, whereas red indicates the incorrect prediction by the LLM. Only 18 (12%) out of 150 assessments were incorrectly predicted, with 132 (88%) correct matching predictions.

**Figure 3.**
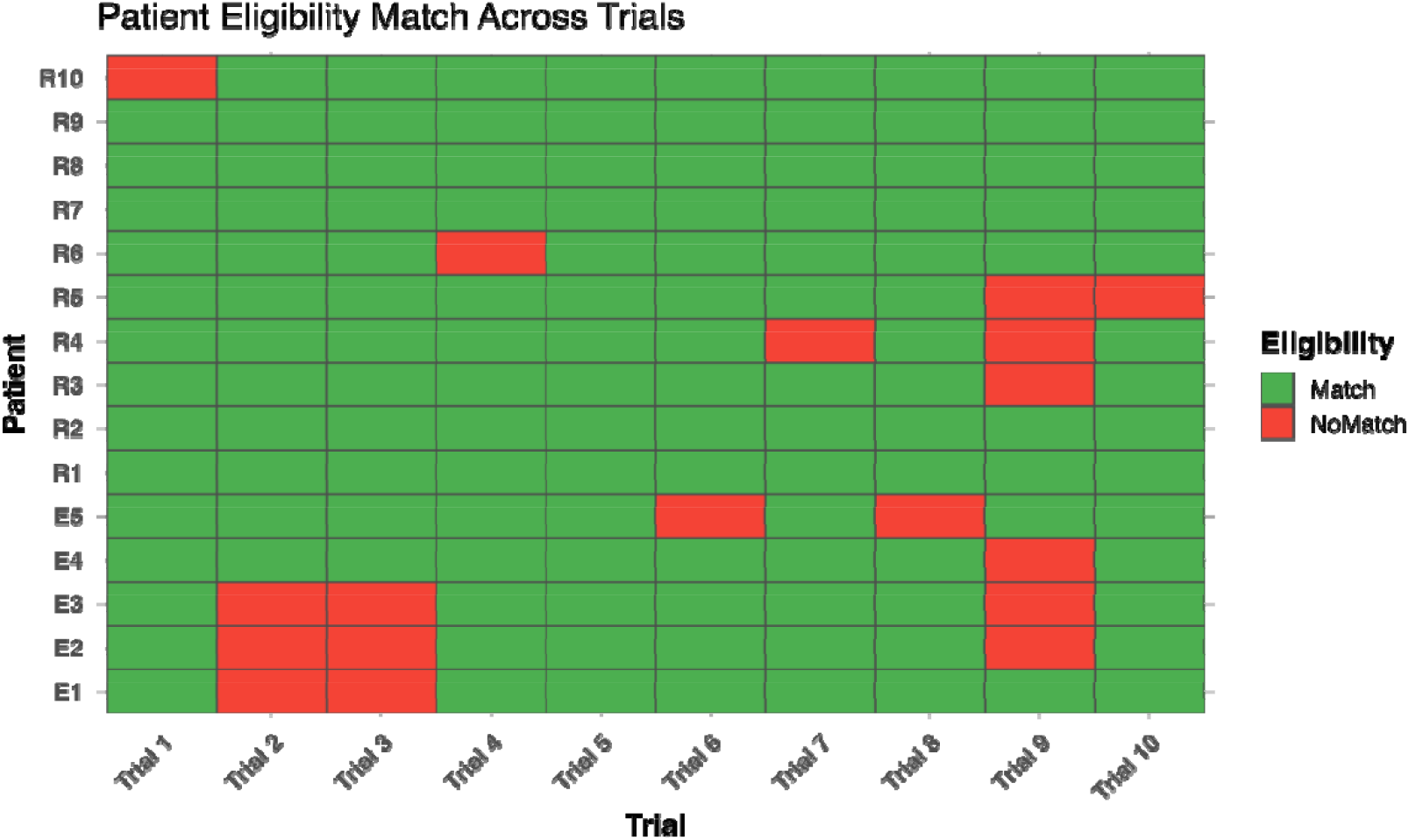
Heatmap representation of the LLM’s eligibility classification across all the trials and the patients.

The average time taken for each trial to evaluate 15 patients shows a clear variability in the performance, as tabulated in Table 2, visually represented in Figure 4. As observed, the LLM takes an average time between 0.40 and 0.50 minutes, significantly lower than the oncologist evaluation of 12.6 and 24.9.

**Table 2:** Patient-Trial evaluation time calculated between Oncologist and LLM (Average per patient in minutes)

| **Trial/Evaluation Time**<br>**(Average in Minutes)** | **Oncologist** | **LLM** |
| --- | --- | --- |
| Trial 1 | 18 | 0.44 |
| Trial 2 | 14.8 | 0.40 |
| Trial 3 | 18.7 | 0.46 |
| Trial 4 | 12.3 | 0.42 |
| Trial 5 | 12.6 | 0.44 |
| Trial 6 | 20 | 0.48 |
| Trial 7 | 15 | 0.46 |
| Trial 8 | 18 | 0.50 |
| Trial 9 | 24.9 | 0.50 |
| Trial 10 | 21.8 | 0.49 |

**Figure 4.**
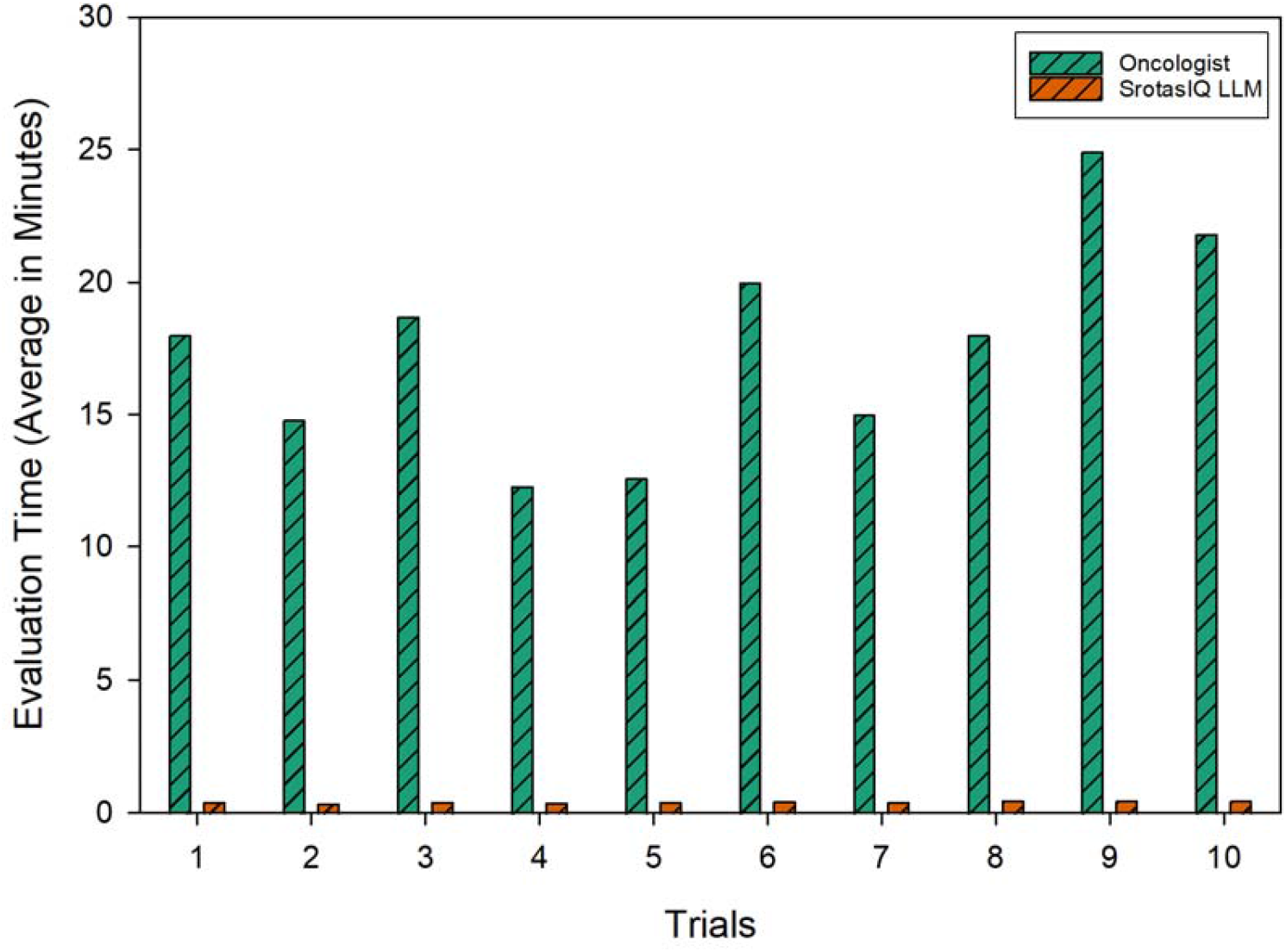
Average patient eligibility assessment across all the trials (oncologist vs SrotasIQ LLM) in minutes.

## 4. Discussion

Automated clinical trial matching systems represent a promising solution to persistent recruitment challenges in oncology. This study validates SROTAS IQ, a domain-specific LLM, using synthetic breast cancer data and expert oncologist review. The controlled evaluation framework enabled systematic assessment across diverse trial types while maintaining clinical authenticity.

Ground truth was established through dual expert review, acknowledging the inherent ambiguity in eligibility decisions. Borderline cases often involve nuanced interpretation of criteria or differing clinical judgments. Our validation framework accounted for this variability, providing a realistic benchmark for automated systems.

Beyond accuracy metrics, we evaluated the clinical coherence of SROTAS IQ explanations. Expert reviewers found the model’s reasoning transparent and useful, particularly in uncertain cases. This supports the importance of explainable AI in clinical settings, where trust and interpretability are essential for adoption.

SROTAS IQ demonstrated commendable performance, with eligibility match rates ranging from 60% to 100% across trials. Several trials achieved near-perfect accuracy, while others highlighted areas for improvement in complex eligibility scenarios.

Time-to-verdict analysis revealed that the LLM completed assessments in under 0.5 minutes per patient, compared to 12–25 minutes for oncologists. While synthetic summaries reduced clinician workload, the LLM’s speed and consistency suggest meaningful efficiency gains. However, this comparison is constrained by the study design: clinicians reviewed concise, pre-assembled summaries rather than navigating fragmented EHR systems. Real-world time savings may vary.

The model’s limitations were most evident in trials with nested sub-criteria, logical dependencies, and domain-specific jargon. These edge cases challenge even experienced clinicians and require deeper contextual understanding. Improving LLM performance in such scenarios will require expanded training datasets with greater heterogeneity and complexity.

Infrastructure demands also warrant consideration. Unlike rule-based systems, LLMs require substantial computational resources for training and inference. Token consumption, GPU availability, and deployment costs must be balanced against clinical utility. Resource optimization strategies will be essential for scalable implementation.

Overall, this study demonstrates the feasibility of LLM-based trial matching in oncology and provides a foundation for future real-world validation. SROTAS IQ shows promise as a decision-support tool, capable of accelerating trial recruitment and reducing administrative burden. Further research should explore generalisability across cancer types, integration into clinical workflows, and health economic impact.

## 5. Conclusions

This study demonstrates a rigorous evaluation of an LLM-based clinical trial matching system in breast cancer using synthetic patient data. The domain-specific model delivered promising results comparable to clinical experts, indicating that automated matching could enhance efficiency and reduce clinical workload. These findings support advancing to translational studies in a real-world populations and underscore the promise of tailored large language models in clinical research, while highlighting key areas for further refinement.

## 6. Limitations and Future Work

This research study has several limitations. First, evaluation was limited to breast cancer and its subtypes, which inherently limits the generalisability of the model to other cancer types and clinical conditions. Second, the study used exclusively synthetic patient data, which may not capture the full complexity and variability of real-world clinical documentation. Third, the validation dataset was relatively small (150 patient-trial combinations), which may limit the robustness of performance estimates across diverse clinical scenarios.

Future work should include real-world validation studies using actual breast cancer patient data and potential expansion to additional cancer types. Prospective assessments could measure effects on trial recruitment efficiency and clinical workflow integration. Additionally, health economic evaluations may help quantify the potential benefits of improved trial matching processes.

## Data Availability

All data produced in the present study are available upon reasonable request to the authors

