## Supplementary File 1 for "SROTAS IQ: An AI-Based Clinical Trial Matching Platform: A Validation Study in Breast Cancer"

### **PROMPT**: "Please generate the following: 15 synthetic breast cancer patients (10 realistic + 5 enriched) for the following trial:

### NCT04227327 (HERMIONE-7) - Study Evaluating Abemaciclib + Aromatase Inhibitors in HR+, HER2- Advanced Breast Cancer Patients (Phase 2)

### INCLUSION CRITERIA:

### Post menopausal women with core biopsy-proven hormone receptor positive invasive breast cancer. Postmenopausal is defined as a woman aged ≥50 years fulfilling any one of the following criteria: i) with amenorrhoea >12 months and an intact uterus; ii) has undergone a bilateral oophorectomy; iii) in women who have undergone a hysterectomy, then FSH levels within the postmenopausal range (utilising ranges from the testing laboratory facility) are required if the patient is aged <55 years; or iv) in women who have been on HRT within the last 12 months and therefore not amenorrhoeic, FSH levels within the postmenopausal range (utilising ranges from the testing laboratory facility) are required if the patient is aged <55 years.

### No evidence of metastatic spread by standard assessment according to local guidelines

### Standard adjuvant endocrine therapy indicated

### A palpable tumour of any size, or a tumour with an ultrasound size of at least 1.5cm

### WHO performance status of 0 or 1

### Written informed consent to participate in the trial and to donation of tissue (fresh tissue and surplus tissue from diagnostic procedures) and blood samples.

### EXCLUSION CRITERIA:

### Locally advanced/inoperable breast cancer

### Evidence of metastatic disease

### Previous invasive breast cancer (surgically treated DCIS or LCIS allowed)

### Current bilateral breast cancer

### Multiple unilateral tumours with different ER/PgR/HER2 status, grade or type (e.g. ductal vs lobular) i.e. anything that suggests two or more different cancers. Multifocal disease with homogenous ER/PgR/HER2 status, grade and type is allowed if at least one lesion is palpable or at least 1.5cm on ultrasound; the largest lesion should be used for sample collection and CRF completion.

### Concurrent use (defined as use within 4 weeks prior to diagnostic tissue sample being taken) of HRT or any other oestrogen-containing medication (including vaginal oestrogens)

### Previous use of oestrogen implants at ANY time

### Prior endocrine therapy or chemotherapy for breast cancer

### Any invasive malignancy diagnosed within previous 5 years (other than basal cell carcinoma or cervical carcinoma in situ)

### Any severe co-incident medical disease, inability to give informed consent or unavailability for follow-up

### Treatment with an unlicensed or investigational drug within 4 weeks before randomisation

### Current, continuous, long term systemic steroid usage

### Requirements:

### 1. No eligibility determinations - Generate patients with detailed clinical information but do not assess whether they meet/don't meet criteria

### 2. Comprehensive clinical detail including:

### Complete staging workup with specific imaging findings

### Detailed biomarker results with scoring (Allred, H-score, etc.)

### Complete histopathology with Nottingham grading components where applicable

### Laboratory values demonstrating organ function

### ECOG performance status and functional assessment

### Complete medical history including prior treatments, surgeries, medications

### Infectious disease screening results

### Cardiac assessment where relevant

### Contraindications and allergy history

### 3. Realistic Population (10 patients): The realistic population was designed to mirror real-world clinical trial screening scenarios, including patients with common exclusion criteria such as prior treatments, comorbidities, premenopausal status, or metastatic disease - reflecting the typical mix of potentially eligible and ineligible cases encountered during actual trial screening.

### 4. Enriched Population (5 patients): The enriched population focused on patients more likely to meet inclusion criteria while varying across clinically relevant dimensions (age, tumor characteristics, staging, etc.), providing adequate representation of the target study population.

### 5. Trial-specific documentation: Include detailed information relevant to the specific inclusion/exclusion criteria

### 6. Clinical authenticity: Each patient should represent realistic clinical scenarios with internally consistent medical histories and findings"

#### **Patient R1: Generated using scenario "Postmenopausal woman with large palpable HR+ breast cancer"**

**Demographic Details:**

- Name: Margaret Thompson
- Age: 67 years
- Gender: Female
- Risk History: Natural menopause at age 52, three children born between ages 24-30, no family history of breast/ovarian cancer

**Medical Timeline:**

**Initial Presentation and Detection Method:** Margaret discovered a large, firm lump in her right breast during self-examination that had been gradually increasing in size over the past 4 months.

**Menstrual/Reproductive History:** Regular menstrual cycles until natural menopause at age 52, no spontaneous menses for 15 years. Three full-term pregnancies with deliveries at ages 24, 27, and 30, breastfed all children for 6-8 months each. No hormone replacement therapy use.

**Clinical Exams and Vitals:** BP 142/86 mmHg, HR 74 bpm, Temperature 98.1°F, Weight 76kg, BMI 28.3. WHO performance status 1. Physical examination revealed a 4.2cm firm, irregular, mobile mass in the upper outer quadrant of the right breast with subtle skin dimpling. Two palpable axillary lymph nodes measuring approximately 1.5cm and 1.2cm.

**Imaging Results:**

- **Mammography:** 4.4cm spiculated mass in right breast upper outer quadrant, BIRADS 5, with associated architectural distortion and pleomorphic calcifications
- **Ultrasound:** 4.1cm irregular hypoechoic mass with angular margins and posterior acoustic shadowing, increased vascularity on Doppler, two suspicious axillary lymph nodes with cortical thickening and loss of fatty hilum
- **Breast MRI:** 4.5cm heterogeneously enhancing lesion with rapid initial enhancement and plateau kinetics, correlating axillary lymph node enhancement, no additional suspicious lesions identified
- **CT Chest/Abdomen/Pelvis:** No evidence of distant metastases, normal liver enhancement, clear lung fields, no suspicious bone lesions or lymphadenopathy beyond regional nodes

**Tumor Characteristics and Staging:** Clinical staging based on imaging and physical examination: cT2N1M0 (Clinical Stage IIB)

**Biomarkers (Central Laboratory Confirmed):**

- **ER:** Positive (94%, Allred score 8, H-score 282, strong nuclear staining in >90% of tumor cells)
- **PR:** Positive (87%, Allred score 7, H-score 261, strong nuclear staining)
- **HER2:** Negative (IHC 0, no membrane staining observed, FISH not performed due to IHC 0 result)
- **Ki-67:** 24% (moderate proliferative activity)

**Detailed Histopathology:**

- **Histologic Type:** Invasive ductal carcinoma, no special type (NST)
- **Grade:** Grade 2 (Nottingham Combined Histologic Grade)
  - **Tubule Formation:** Score 2 (moderate tubule formation, 10-75% of tumor area)
  - **Nuclear Pleomorphism:** Score 2 (moderate nuclear pleomorphism)
  - **Mitotic Count:** Score 2 (12 mitoses per 10 consecutive high-power fields)
  - **Total Score:** 6 (Grade 2)
- **Architecture:** Mixed glandular and solid growth patterns with moderate differentiation
- **Nuclear Features:** Moderate nuclear pleomorphism with nuclei 2-3 times normal size
- **Mitotic Activity:** Moderate mitotic rate (12 mitoses/10 HPF)

**Biopsy Results:** Ultrasound-guided 14-gauge core needle biopsy (8 cores obtained) revealed invasive ductal carcinoma with above characteristics. Image-guided FNA of the larger axillary lymph node showed metastatic adenocarcinoma consistent with breast primary.

**Laboratory Tests (Comprehensive Assessment):**

- **Complete Blood Count:** Hemoglobin 12.8 g/dL, WBC 6.4 K/μL, ANC 3.8 K/μL, Platelets 267 K/μL
- **Comprehensive Metabolic Panel:** Sodium 140 mmol/L, Potassium 4.1 mmol/L, Chloride 102 mmol/L, CO2 24 mmol/L, BUN 22 mg/dL, Creatinine 1.1 mg/dL (eGFR 65 mL/min/1.73m²)
- **Liver Function Tests:** ALT 34 U/L, AST 38 U/L, Alkaline phosphatase 94 U/L, Total bilirubin 0.9 mg/dL, Albumin 4.0 g/dL
- **Hormonal Status Confirmation:** FSH 52 mIU/mL, Estradiol <15 pg/mL (confirming robust postmenopausal status)
- **Coagulation Studies:** PT/INR 1.1, PTT 32 seconds

**Prior Medical History:** Hypertension diagnosed 8 years ago, well-controlled with ACE inhibitor. Hyperlipidemia managed with statin therapy. No history of cancer, autoimmune diseases, or significant surgical procedures.

**Current Medications:** Lisinopril 10mg daily for hypertension, atorvastatin 20mg daily for hyperlipidemia, calcium carbonate with vitamin D3 twice daily, multivitamin. No hormone replacement therapy or estrogenic medications.

**Allergies and Contraindications:** Allergic to penicillin (urticaria). No known contraindications to CDK4/6 inhibitors or aromatase inhibitors.

**Surgical History:** No prior breast surgery, excisional biopsies, or axillary procedures. Cholecystectomy performed 12 years ago without complications.

**Performance Status and Functional Assessment:** WHO Performance Status 1: Restricted in physically strenuous activity but ambulatory and able to carry out work of a light or sedentary nature. Some limitation due to osteoarthritis in knees but independent in all activities of daily living.

**Cardiovascular Assessment:** No history of cardiac disease. Baseline ECG shows normal sinus rhythm. Blood pressure well-controlled on current antihypertensive therapy.

**Comorbidities:** Mild osteoarthritis affecting knees and hands, managed with occasional acetaminophen. Osteopenia on recent DEXA scan (T-score -1.8 lumbar spine).

**Treatment History:** No prior chemotherapy, endocrine therapy, or radiation therapy. No previous treatment for breast cancer or other malignancies.
