## Supplementary File 2 for "SROTAS IQ: An AI-Based Clinical Trial Matching Platform: A Validation Study in Breast Cancer"

**Actual Table (Average Score among 15 Patients):**

| **Trials** | **Accuracy** | **Precision** | **Recall** | **Specificity** | **F1-Score** | **PPV** | **NPV** |
| --- | --- | --- | --- | --- | --- | --- | --- |
| **Trial 1** | 97.79 | 95.83 | 97.35 | 96.65 | 96.37 | 95.83 | 97.77 |
| **Trial 2** | 92.96 | 79.47 | 88.11 | 94.13 | 81.21 | 79.47 | 97.32 |
| **Trial 3** | 86.38 | 79.99 | 80.85 | 89.32 | 79.69 | 79.99 | 90.23 |
| **Trial 4** | 98.14 | 97.60 | 98.56 | 97.82 | 98.04 | 97.60 | 98.38 |
| **Trial 5** | 75.55 | 65.15 | 95.66 | 61.17 | 76.23 | 65.15 | 96.16 |
| **Trial 6** | 85.95 | 70.53 | 94.77 | 84.70 | 74.82 | 70.53 | 99.17 |
| **Trial 7** | 93.33 | 86.83 | 97.33 | 91.36 | 90.97 | 86.83 | 98.93 |
| **Trial 8** | 92.63 | 87.47 | 96.66 | 89.76 | 91.60 | 87.47 | 97.54 |
| **Trial 9** | 80.83 | 70.10 | 53.83 | 92.81 | 58.91 | 70.10 | 82.89 |
| **Trial 10** | 86.19 | 81.03 | 87.12 | 85.80 | 82.56 | 81.03 | 92.98 |

**Normalised Table (min-max norm):**

| **Trials** | **Accuracy** | **Precision** | **Recall** | **Specificity** | **F1-Score** | **PPV** | **NPV** |
| --- | --- | --- | --- | --- | --- | --- | --- |
| **Trial 1** | 0.9845064 | 0.9454545 | 0.9729488 | 0.9680764 | 0.9573217 | 0.9454545 | 0.9140049 |
| **Trial 2** | 0.770695 | 0.4412943 | 0.7663760 | 0.8993179 | 0.5698952 | 0.4412943 | 0.8863636 |
| **Trial 3** | 0.4794157 | 0.457319 | 0.6040689 | 0.7680764 | 0.5310503 | 0.457319 | 0.4508600 |
| **Trial 4** | 1 | 1 | 1.0000000 | 1 | 1 | 1 | 0.9514742 |
| **Trial 5** | 0 | 0 | 0.9351666 | 0 | 0.4426271 | 0 | 0.8151106 |
| **Trial 6** | 0.4603807 | 0.1657935 | 0.9152694 | 0.6420191 | 0.4065934 | 0.1657935 | 1.0000000 |
| **Trial 7** | 0.7870739 | 0.6681048 | 0.9725017 | 0.8237381 | 0.8193202 | 0.6681048 | 0.9852580 |
| **Trial 8** | 0.7560868 | 0.6878274 | 0.9575229 | 0.7800819 | 0.8354204 | 0.6878274 | 0.8998771 |
| **Trial 9** | 0.2337317 | 0.1525424 | 0.0000000 | 0.8633015 | 0 | 0.1525424 | 0.0000000 |
| **Trial 10** | 0.4710049 | 0.4893683 | 0.7442432 | 0.6720327 | 0.6043956 | 0.4893683 | 0.6197789 |
